# A Proof-of-Concept Large Language Model Application to Support Clinical Trial Screening in Surgical Oncology

**DOI:** 10.1101/2024.09.20.24314053

**Authors:** Samantha M. Lai, Alysala M. Malik, Tejas S. Sathe, Caitlin J. Silvestri, Gulam A. Manji, Michael D. Kluger

## Abstract

**Introduction:** Clinical trials advance the forefront of medical knowledge and rely on consistent patient accrual for success. However, patient screening for clinical trials is resource intensive. There is a need to increase the scalability of trial recruitment while maintaining or improving upon the sensitivity of the current process. We hypothesized we could use a state-of-the-art large language model (LLM), prompt engineering, and publicly available clinical trial data to predict patient eligibility for trials from clinic notes. Here, we present pilot data demonstrating the accuracy of this tool in a cohort of patients being evaluated for pancreas cancer treatment.

**Methods:** Patients who were screened for clinical trials at a single institution were studied. An LLM application was developed using LangChain and the GPT-4o model to assist in clinical trial screening. Deidentified patient data from clinical notes and trial eligibility criteria from ClinicalTrials.gov were used as inputs. For each patient, the model determined inclusion or exclusion with respect to selected eligibility criteria as well as nine clinical trials. Model responses were graded programmatically against a human rater standard. Time elapsed and cost for running each analysis were recorded.

**Results:** Of the 24 patients in the test set, 19 were eligible for at least one trial. There were 43 eligible patient-trial matches in the data set. Our model correctly predicted 39 out of 42 (90.7%) of these matches. There were 105 individual eligibility criteria evaluated per patient for a total of 2520 binary criteria. GPT-4o agreed with the raters for 2,438 out of 2,520 (96.7%) binary eligibility criteria. Sensitivity to overall trial eligibility ranged from 87.5% to 100% for 8 out of 9 trials. Specificity ranged from 73.3% to 100% over all nine trials. The median cost for screening a patient was 0.67 USD (0.63-0.74). Median time elapsed was 137.66 seconds (130.04-146.04). Median total token usage across three assistants was 112,266.5 tokens (102,982.0-122,174.2).

**Conclusion:** Overall, this model showed high sensitivity and specificity in using minimally processed free-text clinical notes to screen patients for appropriate clinical trials using a fraction of the time and cost of existing screening mechanisms. Results showed promise with a small cohort, and future studies are needed to assess its accuracy with a larger sample of patients and trials. This study represents the frontier of pitting of emerging large language model technology against the historically unruly terrain of the electronic medical record, suggesting that the imperfection of free-text clinical notes only slightly hinders the performance of a general-use model compared to previous performance on preprocessed data. These findings highlight that using this tool directly on clinical notes could complement human screening efforts to improve patient accrual at a low time and monetary cost.

## Introduction

Clinical trials advance the forefront of medical knowledge and rely on consistent patient accrual for success. Patient screening for clinical trials is a resource intensive process that requires cross-referencing vast amounts of clinical information in patient charts against specific trial eligibility criteria. Finding, screening, and enrolling a patient can take up to eight hours and cost hundreds of dollars depending on the phase of the trial.^1^ Screening often occurs in an ad hoc manner, ranging from physician discussion at interdisciplinary tumor boards to a dedicated clinical screener splitting time between recruiting patients over the phone and searching for potentially eligible patients in the electronic medical record. Fragmented efforts such as these may contribute to the low rate of enrollment in cancer trials in the United States.^2^ Other consequences of inefficiencies and inaccuracies range from limiting a patient’s access to the newest medical advances to the significant economic burden felt by academic medical centers suffering from low accrual and subsequent failed trials.^1,3,4^ There is room to grow both in terms of improving patient access to trials and reducing waste in the system at large.^3,5^ This calls for increased scalability of trial recruitment while maintaining the sensitivity of the current process.

Generative artificial intelligence is an emerging technology that shows promise in meeting this need. Large language models (LLMs) can synthesize clinical information as well as capture information from unstructured clinical materials such as notes and imaging reports.^6^ They have been shown to use preprocessed free text clinical data to glean the individual criteria necessary to determine eligibility and reason about an overall clinical picture as it relates to eligibility. However, an LLM has not yet been shown to effectively screen a patient for both individual eligibility criteria and overall trial eligibility using minimally-processed, free-text clinical notes and unstructured eligibility criteria.^7–9^ This gap results from the limited size of the context window of many general-use LLMs as well as patient privacy protections limiting the transmission of protected health information through proprietary application programmer interfaces (APIs).^7^

We hypothesized we could use a state-of-the-art LLM, prompt engineering, and publicly available clinical trial data to predict patient eligibility for clinical trials from clinic notes. Here, we present pilot data demonstrating the accuracy of our tool in a cohort of patients being evaluated for pancreas cancer treatment.

## Methods

### Chart review

We identified patients who were screened for enrollment in clinical trials at the Pancreas Center between January and May 2024. Thirty-two patients who ultimately pursued treatment at the center were identified for further study. All nine ongoing trials at the Pancreas Center were used for this study. Eight patient charts representing eight of the nine trials were set aside as training data for developing the prompts, leaving 24 patients in the test set. Characteristics of the patients in the training and test sets are available in Table S1. A medical oncology progress note was identified in the medical record. If available, a surgical oncology note was also utilized. All identifying information was removed manually by study personnel in accordance with the Safe Harbor method for deidentification.^10^ Additionally, any references to specific clinical trials by an identifiable name or by NCT number were removed.

### Two-rater Standard for Eligibility Criteria Review

Eligibility criteria were retrieved from ClinicalTrials.gov for each trial using the trial NCT number and deconstructed into one or multiple binary criteria that, taken together, captured the information necessary to determine whether the patient met the ClinicalTrials.gov criterion (Figure 1a). This process resulted in 96 unique binary eligibility criteria. The deidentified notes were then reviewed by two independent raters to determine the patient’s status with respect to the 96 binary criteria and eligibility for 9 clinical trials for a total of 105 variables (Table S2a). Eligibility for inclusion in a trial was defined as meeting all binary inclusion criteria specific to the trial while not meeting any exclusion criteria specific to the trial. Raters were trained in clinical trials screening but were not board-certified medical or surgical oncologists. All inter-rater discrepancies in answers were resolved verbally between the raters.

**Figure 1a.**
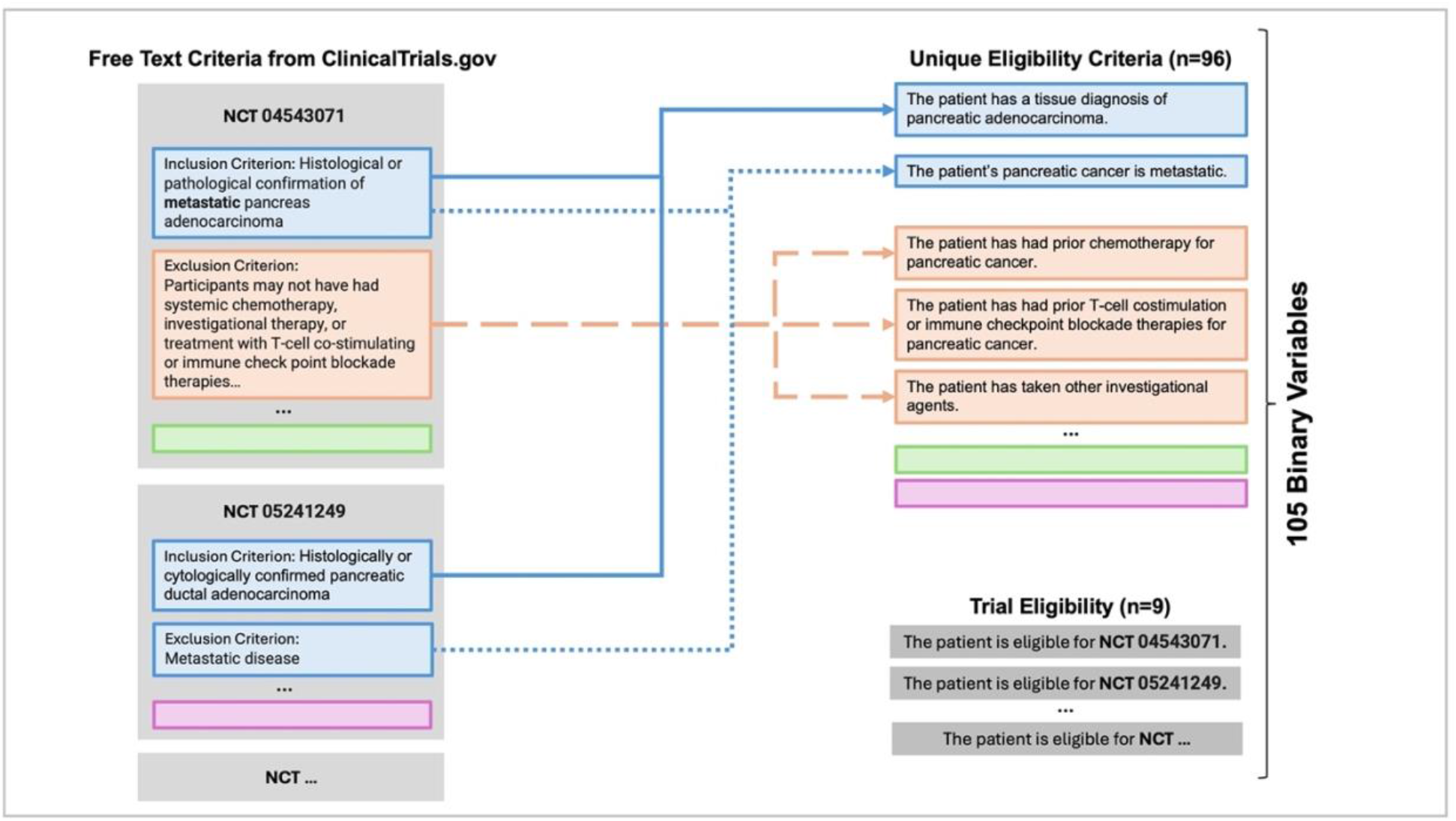
Construction of Unique Binary Variables This depicts the process of creating a total of 105 binary statements out of the free-text trial inclusion and exclusion criteria found on ClinicalTrials.gov.

**Figure 1b.**
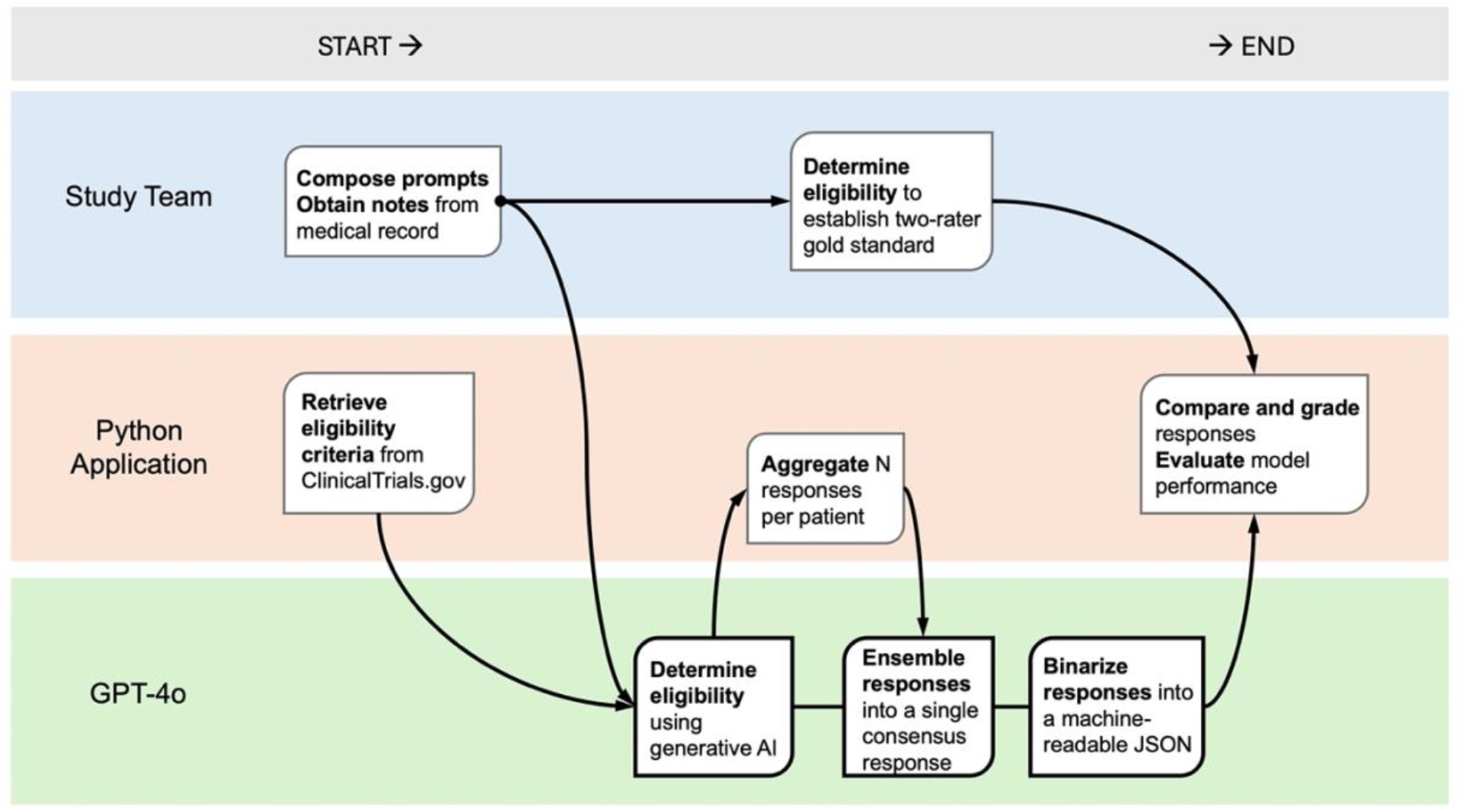
Depiction of Data Flow, Inputs, and Outputs The workflow depicts the flow of data from preprocessing of clinical notes by study personnel and automated retrieval of eligibility criteria from the ClinicalTrials.gov API. GPT-4o denotes Generative Pre-trained Transformer 4 Omni; AI, artificial intelligence; JSON, JavaScript object notation.

### Application Structure

An LLM application using the Python LangChain library and the OpenAI Generative Pre-trained Transformer 4 Omni (GPT-4o) model with all default configuration settings was developed (Figure 1).^11,12^ The application consisted of three specialized chains, or sequences of action, each of which invoked the LLM with a different prompt. Cost of the OpenAI API calls and time elapsed per patient were measured programmatically.

The first chain, called the eligibility chain, received two items as input: the deidentified notes from one patient and the eligibility criteria for all the clinical trials. The eligibility criteria were retrieved by NCT number using the ClinicalTrials.gov API. The chain was then prompted to evaluate each of the 105 binary eligibility criteria by providing the patient’s eligibility status with respect to each criterion or trial and a brief sentence to explain each of its answers (Table S2a).

The eligibility screening process was run five times for each patient, resulting in five separate answers per patient. A second chain, called the ensembling chain, received the five answers for each binary criterion and derived a consensus answer by consolidating the five answers based on the majority answer to reduce the effect of any single hallucination and improve the reliability of the LLM output as suggested by Nori et al in the development of MedPrompt.

A final chain, called the binarization chain, was prompted to receive the output of the ensembling chain and build a machine-readable JSON (JavaScript Object Notation) in a standardized format specified in the prompt (Table S2b). Prompts did not contain input and output examples, relying instead on the zero-shot learning capabilities of GPT-4o. Prompts were iteratively adjusted based on mistakes made on the test set until trial eligibility performance on the test set reached a plateau.

### Grading

The JSON objects outputted by the LLM application were graded programmatically by a Python grading function comparing the model output to the two-rater gold standard. True positive and true negatives were respectively defined as identifying eligibility or ineligibility for a trial in alignment with a two-rater gold standard. Sensitivity and specificity were calculated for each criterion and overall trial eligibility. Discrepancies between the LLM application and the inter-rater gold standard were further categorized by the raters into several types of errors. Error category definitions and discussion on specific errors are included in Supplementary Materials. Examples of the errors are detailed in the Results section.

## Results

### Performance on Trial Eligibility and Individual Criteria

Of the 24 patients in the test set, 19 were eligible for at least one trial. The model correctly predicted 39 out of 42 (90.7%) of eligible patient-trial matches in the data set. There were 105 individual eligibility criteria evaluated per patient for a total of 2,520 binary criteria. GPT-4o agreed with the raters for 2,438 out of 2,520 (96.7%) binary eligibility criteria.

Sensitivity to overall trial eligibility by trial ranged from 87.5% to 100% for 8 out of 9 trials. The application incorrectly screened the single patient in one trial test set, leading to a sensitivity of 0% for this trial. Specificity ranged from 73.3% to 100% over all nine trials (Figure 2). Performance of all individual criteria are included in Table S3.

**Figure 2.**
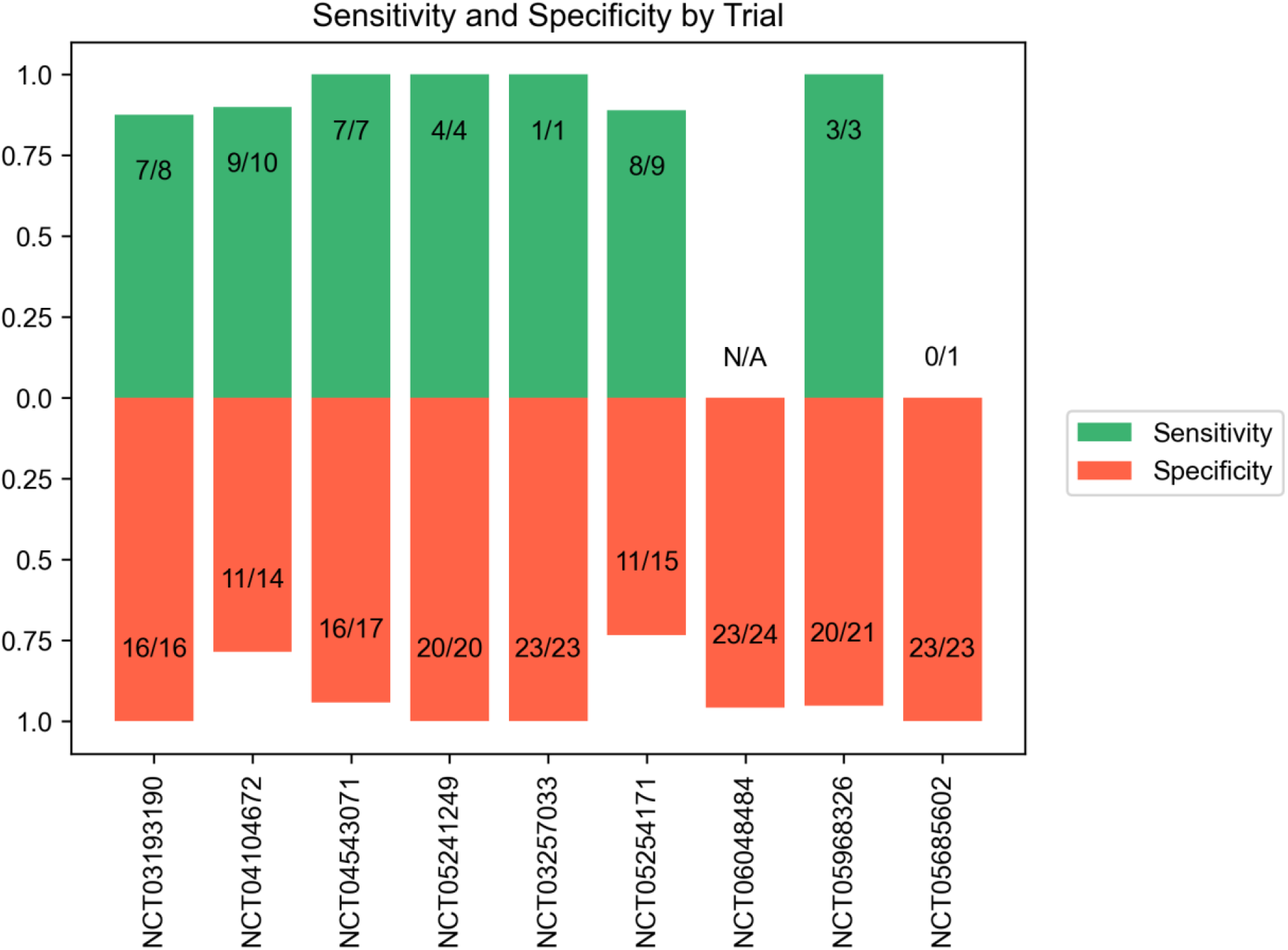
Sensitivity and Specificity for Trial Eligibility This data represents a single experiment run on September 9, 2024. Each pair of green and orange bars represents sensitivity and specificity for GPT-4o determination of trial eligibility across nine trials for pancreatic cancer, with each trial denoted by the National Clinical Trial (NCT) number on the X axis. The number of patients included in each calculation and odds ratios are indicated as N(predicted)/N(total).

### Cost and Time Elapsed

The median cost for screening a patient was 0.67 USD (0.63-0.74). Median time elapsed was 138 seconds (130-146). Median total token usage across three assistants was 112,266.5 tokens (102982.0-122174.2), with most tokens used in the eligibility determination stage. The median token usage in this part of the application was 96571.5 (87497.5-106799.0), well within GPT-4o’s context window of 120,000 tokens. Further breakdown of resource usage is detailed in Table S4.

### Error Analysis

Out of 2520 variables evaluated in total across 24 patients encompassing individual eligibility criteria and trial eligibility, 82 discrepancies were found upon grading. The most common source of discrepancy was human error in chart review. Examples of each error category are provided in Table 2 and a breakdown of the variables in each error category is presented in Figure 3. Further discussion of errors in included in Supplementary Materials.

**Table 1.** Selected Individual Eligibility Criteria. This data represents a single experiment run on September 9, 2024. Each row represents sensitivity and specificity for GPT-4o determination of individual criterion status. The number of patients included in each calculation is indicated.

| Criterion | Sensitivity (TP for exclusion) | Sensitivity Calculation | Specificity (TN for inclusion) | Specificity Calculation |
| --- | --- | --- | --- | --- |
| The patient's age is less than 18. | 0.0 | N/A | 1.000000 | 24/24 |
| The patient's pancreatic cancer is resectable. | 1.0 | 5/5 | 1.000000 | 19/19 |
| The patient's pancreatic cancer is metastatic. | 1.0 | 15/15 | 1.000000 | 9/9 |
| The patient has had disease progression on 5-FU for metastatic or unresectable pancreatic ductal adenocarcinoma. | 0.0 | 0/1 | 1.000000 | 23/23 |
| ECOG performance status (PS) is greater than 2. | 0.5 | 1/2 | 1.000000 | 22/22 |
| The patient is taking metformin. | 0.2 | 1/5 | 0.947368 | 18/19 |
| The patient's total bilirubin is >1.5x upper limit of normal or >3x upper limit or normal in patients with Gilbert disease | 0.5 | 1/2 | 0.954545 | 21/22 |
| The patient has active or symptomatic Hepatitis B | 0.0 | 0/1 | 1.000000 | 23/23 |

**Table 2.** Discrepancies between GPT-4o and Human determinations of individual eligibility criteria. A total of 82 discrepancies were categorized after programmatic grading. The number and percentage of the total number of errors is represented. A true positive is denoted when the criterion statement is true and the rater or model answers as such. GPT-4o denotes Generative Pre-trained Transformer 4 Omni; GI, gastrointestinal; 5-FU, 5-fluorouracil.

| Category | Criterion | Correctness of GPT-4o and Human Answers | Explanation | N (%) |
| --- | --- | --- | --- | --- |
| Prompt Unclear | The patient has a diagnosis of pancreatic adenocarcinoma. | GPT-4o False Positive, Human True Negative | GPT-4o states there is a diagnosis of "pancreatic adenocarcinoma", but it is signet cell based, not pancreatic ductal adenocarcinoma. This was an error of prompt engineering. | 2 (2.4) |
| Note Unclear | The patient has a history of other malignancy within 2 years that does not include malignancies with a low rate of metastatic spread. | GPT-4o False Positive, Human True Negative | Patient had renal cell carcinoma, but it was resected in 2015. The model had no way of knowing the date of the note. By study design, all patients were all seen within the last year, so we had information the model was unable to access in the deidentified note. | 5 (6.1) |
| Clinical Judgment | The patient has a history of GI condition which could impair absorption or ability to ingest study drug (i.e. evidence of GI obstruction). | GPT-4o False Positive, Human True Negative | Patient is status post duodenal stent placement. GPT-4o did not connect duodenal stent placement mentioned in the note to the small bowel obstruction. | 12 (14.6) |
| Misinterpretation | The patient has had disease progression on 5-FU for metastatic or unresectable pancreatic ductal adenocarcinoma. | GPT-4o False Negative, Human True Positive | GPT-4o was unable to identify that the patient received 5-FU for metastatic disease and experienced disease progression despite the treatment. | 14 (17.0) |
| Misinterpretation | The patient is eligible for trial NCT04543071. | GPT-4o False Positive, Human True Negative | GPT-4o screened the patient into this trial. However, the patient must be naïve to treatment for metastatic disease for this trial. | See above |
| Hallucination | The patient's hemoglobin is less than 9.0 g/dL. | GPT-4o False Positive, Human True Negative | The note states in the laboratory values section that the hemoglobin is 10.2. | 23 (28.0) |
| Human Error | The patient has had prior radiotherapy for pancreatic cancer. | GPT-4o True Positive, Human False Negative | The note states that the patient had radiotherapy, and this was missed by the raters. | 26 (31.7) |

**Figure 3.**
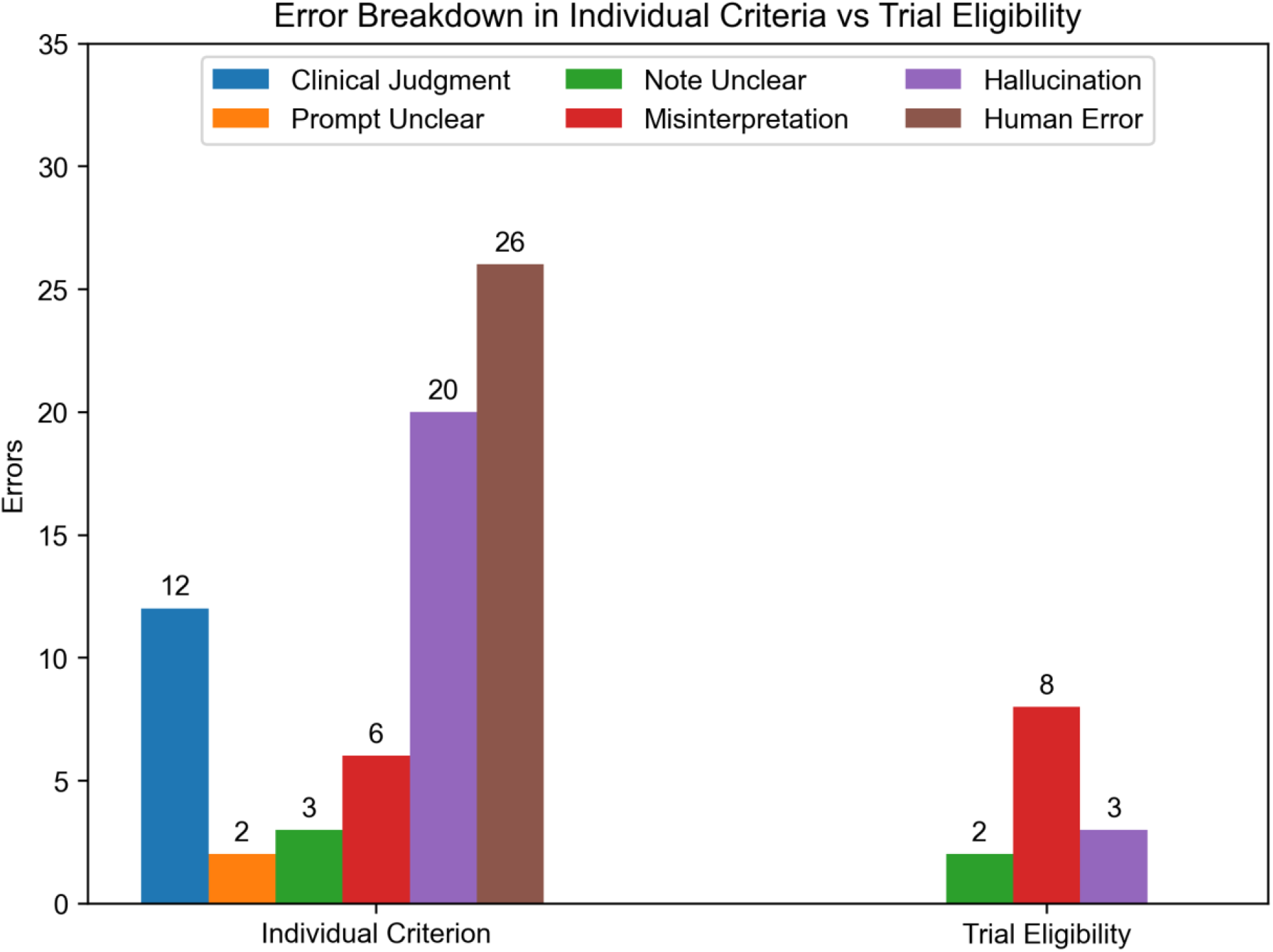
Error Breakdown Errors from one experiment on September 9, 2024 were categorized into six error types and separated into errors in determining individual eligibility criteria and errors in determining overall trial eligibility.

## Discussion

Overall, our application shows high sensitivity and specificity in selecting patients for appropriate clinical trials, using a fraction of the time and cost of existing screening mechanisms.^1^ The application of LLMs to the problem of clinical trials screening has been found to show promise in isolated tasks, such as extraction of individual eligibility criteria; understanding preprocessed clinical summaries and case reports; and evaluation of structured eligibility data.^7–9^ However, a gap in applicability to clinical practice exists because restrictions in context window length and patient privacy protections have prevented studies from evaluating screening capabilities using realistic clinical notes.

In our application, we display a general-use LLM’s ability to use imperfect and unstructured data to make an algorithmic decision. Our findings highlight that the imperfection of free-text clinical notes only slightly hinders the performance of a general-use model when compared to the performance in other studies on more preprocessed data.^7–9^ Simultaneously evaluating both GPT-4o’s ability to extract individual eligibility criteria as well as overall trial eligibility provides a proxy for explainability alongside insight into the strengths and limitations of the technology. For example, GPT-4o’s near perfect performance on criteria such as age and resectability of pancreatic neoplasm show that it can reliably glean these details from a free-text clinical note. In addition to explainability, this data may be useful to physicians in finding alternate trials for which patients are eligible or reconsidering eligibility if any of the “missed” criteria represent clinical conditions that may resolve.

Several limitations were encountered in this study. First, the pool of patients enrolled in trials at a single center for pancreatic cancer was relatively small. In optimizing for performance on enrolled patients at one center, the application may have been overfit to the relatively small cohort, which may limit the generalizability of our results. Future directions will involve validation in larger and more diverse datasets. A limitation inherent to this field is the black box nature of generative AI obscuring explainability, which we attempted to mitigate with chain-of-thought prompting to shed light on basic information that the model could extract in isolation alongside demonstration of higher-level reasoning.

Our findings suggest that GPT-4o’s performance on individual criteria can rival that of trained non-physician screeners and provide a complementary set of strengths in support of a largely human process. These results concur with preprinted findings that state that general-use models can display impressive performance “out of the box” without specialization beyond application-specific prompting. The trial-agnostic design of our application can easily substitute different clinical trials to explore generalizability of the tool. We invite further evaluation in a high-fidelity simulation of the screening process with prospective comparison in resource use, time, and accuracy to current processes. Future validation on different models, both open-source and commercially available, could be easily implemented to explore generalizability, increase feasibility for integration into existing electronic health record systems, and elucidate the upper limit of performance on this important clinical problem.

## Conclusion

This study represents the frontier of pitting of emerging large language model technology against the historically unruly terrain of the electronic medical record, suggesting that the imperfection of free-text clinical notes only slightly hinders the performance of a general-use model compared to previous performance on preprocessed data. These findings highlight that using this tool directly on clinical notes could complement human screening efforts to improve patient accrual at a low time and monetary cost.

## Data Availability

All data produced in the present study are available upon reasonable request to the authors

https://github.com/laisamantha/trial-find

## Acknowledgments

The authors appreciate Justine Lugo for her dedication to patients and for her help in defining the focus of this study. S.L. thanks Peter McEvoy for technical guidance in software development.

## References

1. Penberthy LT, Dahman BA, Petkov VI, DeShazo JP. Effort Required in Eligibility Screening for Clinical Trials. J Oncol Pract. 2012;8(6):365–370. doi:10.1200/JOP.2012.000646

2. Unger JM, Shulman LN, Facktor MA, Nelson H, Fleury ME. National Estimates of the Participation of Patients With Cancer in Clinical Research Studies Based on Commission on Cancer Accreditation Data. J Clin Oncol. Published online April 2, 2024. doi:10.1200/JCO.23.01030

3. Kitterman DR, Cheng SK, Dilts DM, Orwoll ES. The Prevalence and Economic Impact of Low-Enrolling Clinical Studies at an Academic Medical Center. Acad Med J Assoc Am Med Coll. 2011;86(11):1360–1366. doi:10.1097/ACM.0b013e3182306440

4. Woo M. An AI boost for clinical trials. Nature. 2019;573(7775):S100–S102. doi:10.1038/d41586-019-02871-3

5. Sateren WB, Trimble EL, Abrams J, et al. How Sociodemographics, Presence of Oncology Specialists, and Hospital Cancer Programs Affect Accrual to Cancer Treatment Trials. J Clin Oncol. Published online September 21, 2016. doi:10.1200/JCO.2002.08.056

6. Singhal K, Azizi S, Tu T, et al. Large language models encode clinical knowledge. Nature. 2023;620(7972):172–180. doi:10.1038/s41586-023-06291-2

7. Wong C, Zhang S, Gu Y, et al. Scaling Clinical Trial Matching Using Large Language Models: A Case Study in Oncology. In: Proceedings of the 8th Machine Learning for Healthcare Conference. PMLR; 2023:846–862. Accessed September 16, 2024. https://proceedings.mlr.press/v219/wong23a.html

8. Beattie J, Neufeld S, Yang D, et al. Utilizing Large Language Models for Enhanced Clinical Trial Matching: A Study on Automation in Patient Screening. Cureus. 16(5):e60044. doi:10.7759/cureus.60044

9. Unlu O, Shin J, Mailly CJ, et al. Retrieval-Augmented Generation–Enabled GPT-4 for Clinical Trial Screening. NEJM AI. 2024;1(7). doi:10.1056/AIoa2400181

10. Rights (OCR) O for C. Guidance Regarding Methods for De-identification of Protected Health Information in Accordance with the Health Insurance Portability and Accountability Act (HIPAA) Privacy Rule. September 7, 2012. Accessed September 18, 2024. https://www.hhs.gov/hipaa/for-professionals/special-topics/de-identification/index.html

11. Hello GPT-4o. Accessed September 18, 2024. https://openai.com/index/hello-gpt-4o/

12. Introduction | 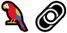 LangChain. Accessed September 18, 2024. https://python.langchain.com/docs/introduction/

