## Supplementary Materials for "A Proof-of-Concept Large Language Model Application to Support Clinical Trial Screening in Surgical Oncology"

Samantha M. Lai, BA

Alysala M. Malik, BA

Tejas S. Sathe, MD

Caitlin J. Silvestri, MD

Gulam A. Manji, MD, PhD

Michael D. Kluger, MD, MPH

### Code availability

Code available on GitHub at: https://github.com/laisamantha/trial-find

### Methods: Error Definitions

A “Human Error” was defined as a discrepancy where the information needed to arrive at the correct answer was in the clinical note and extracted by GPT-4o, but incorrectly annotated by the raters. “Prompt Unclear” and “Note Unclear” were assigned to discrepancies where the information required to answer the criterion was deemed to be ambiguous by the raters. A “Clinical Judgment” error was invoked in cases where GPT-4o differed from the raters in appraisal of a piece of information in the note, such as the severity of a condition. A “Misinterpretation” error was defined as an instance where GPT-4o did not seem capable of extracting the relevant information from the note. A “Hallucination” was defined as an error where the answer provided by GPT-4o actively contradicted information stated in the note.

### Results: Error Analysis

The most common source of discrepancy was human error in chart review. The other most common categories of discrepancies were GPT-4o hallucination, GPT-4o misinterpretation, and disagreements in clinical judgement. Thirteen out of 82 discrepancies were in overall trial eligibility determination, leaving 69 errors in determination of individual criteria. All 26 human errors categorized were entirely in determination of individual criteria, making up 37.7% of individual criterion errors. Hallucination and misinterpretation together accounted for 26 criterion errors, or 37.7% of the total criterion errors. These two categories made up 11 out of 13 (84.6%) of the overall trial eligibility determination errors.

The model generated some errors which preclude immediate widespread use in the clinical setting. For example, one patient was incorrectly deemed eligible for four trials with an eligibility criterion requiring that patients be naïve to treatment for metastatic disease. Sure enough, the individual criterion assessing whether the patient had experienced progression of metastatic disease on chemotherapy was also incorrect, suggesting that the model had also struggled to pick up on a piece of crucial information needed to score correctly on overall eligibility. GPT-4o’s sensitivity for detecting metformin on a patient’s medication list was surprisingly low, possibly because the format in which the medication list differed from the text data on which GPT-4o was trained. Out of five patients taking metformin, GPT-4o was only able to detect one, in contrast to its perfect record on Hepatitis B status or its near-perfect performance on tumor staging, both which appear in a much less standardized form than metformin in typical oncology notes.

### Supplementary Table S1A. Methods: Demographics and Characteristics of the Study Population

| Sample Characteristic | Development Set (n=8) | Test Set (n=24) |
| --- | --- | --- |
| Age, years (Mean [SD]) | 64.8 [11.5] | 64.8 [9.4] |
| Age, years  18-40  41-60  61-80  81+ | 0 (0)  4 (50)  3 (38%)  1 (13%) | 1 (4%)  5 (21%)  18 (75%)  0 (0%) |
| Male Sex | 4 (50%) | 12 (50%) |
| Race  White  Black  Asian  Other  Declined to State | 4 (50%)  0 (0%)  1 (13%)  2 (25%)  1 (13%) | 13 (54%)  3 (12%)  2 (8%)  5 (21%)  1 (4%) |
| Diagnosis and Resectability  Resectable PDAC  Borderline resectable PDAC  Locally advanced PDAC  Metastatic PDAC  Other Diagnosis | 1 (13%)  2 (25%)  1 (12%)  4 (50%)  0 (0%) | 5 (21%)  3 (13%)  1 (4%)  14 (58%)  1 (4%) |

*Values are reported as N (%) unless otherwise noted

**PDAC refers to pancreatic ductal adenocarcinoma

### Supplementary Table S1b. Methods: Representativeness of the Study Population

| Disease/Problem under investigation | Pancreatic cancer | Clinical Trials Screening |
| --- | --- | --- |
| Sex and gender | The incidence of pancreatic cancer is slightly higher in men compared to in women.^1^ | A slight majority of cancer trial patients are female, with no significant difference in accrual between the sexes.^2^ |
| Age | Risk of developing pancreatic cancer increases with age. The most common age group for pancreatic cancer incidence is 55-79 years, accounting for around 50% of cases.^1^ | Pediatric patients are accrued into trials at a high rate, but only a small percentage of adult cancer patients are enrolled into clinical trials.^2^ |
| Race or ethnic group | SEER data from 2011-2015 display that incidence and mortality rates were higher in Black patients than any other racial or ethnic group. Incidence was lowest in AAPI patients.^3^ | A smaller proportion of Black, Hispanic, and Asian American adults are accrued into trials compared to White patients of the same age.^2^ |
| Geography | Higher rates of pancreatic cancer have been observed in North America, with lower rates observed in the Eastern Mediterranean region and Southeast Asia.^3^ | Geographic areas with higher socioeconomic levels experience higher levels of clinical trial accruals.^2^ |
| Other considerations | The rate of diagnosis of pancreatic cancer may be affected by the Human Development Index and health care access, confounding the point above concerning the rate of pancreatic cancer across geographies. Other theories have considered the role of latitude and vitamin D levels in the development of pancreatic cancer.^3^ | Patients enrolled into clinical trials are less likely to be uninsured and more likely to be covered by Medicare.^2^ |
| Overall representativeness | Our study population, which consisted of all patients currently enrolled at the CUIMC Pancreas Center, was predominantly White. Given the epidemiology of pancreatic cancer, we recognize that our findings may not generalize to a more accurate sampling of patients with pancreatic cancer and note that this is a necessary avenue for future work. | Our study population reflects the imbalanced demographics of the data reported above. To accurately gauge our tool’s effectiveness, prospective studies are needed to sample a more diverse and representative population of cancer patients. |

### Supplementary Table 2a. Methods: Eligibility

| **Eligibility** | You are an assistant that helps to screen patients for eligibility for clinical trials for pancreatic adenocarcinoma. You will be provided a free text oncology and/or surgical oncology note about a patient. Pay special attention to whether the patient's disease is resectable, borderline resectable, locally advanced, or metastatic, as these are mtually exclusive. Also pay special attention to what treatments the patient has undergone and for what indication. I will then give you a series of 106 true or false statements.  Patient Clinical Note  {scenario}  End Patient Clinical Note  We will then assess the patient for clinical trial eligibility in a similar way. You will be provided with the Inclusion and Exclusion Criteria for a set of clinical trials, which are identified with unique IDs starting with 'NCT'. If a patient meets some of the Inclusion Criteria but some remain indeterminate due to missing information, they may still be eligible for the study as more information about the patient may be determined later in the screening process. Thus, for a given trial, patients who meet some or all the inclusion criteria and none of the exclusion criteria are considered "Eligible". However, if they meet one or more of the Exclusion Criteria or if one or more Inclusion Criteria are contradicted by the patient note, they are "Not Eligible". In your final determination, patients will either be "Eligible" or "Not Eligible". Detail your reasoning for the patient's eligibility for each of the trials in responses under 30 tokens per trial. Give your answer in the form: "Eligible for trial [NCT number]. [Reasoning]." or "Not Eligible for trial [NCT number]. [Reasoning]."  Clinical Trials Eligibility Criteria  {context}  End Clinical Trials Eligibility Criteria  Here are the true/false statments along with some guidance about how to answer based on the topic based on previous errors you have made. For each statement, please consider the answer guidance and give a concise answer under 20 tokens about what makes the statement true or false. Do not restate the statements below in your answer unless it is about trial eligibility.  1. Age: The patient's age is less than 18.  2. Diagnosis: The patient has a diagnosis of pancreatic adenocarcinoma.  3. Diagnosis: The patient has a diagnosis of pancreatic acinar cell carcinoma.  4. Diagnosis: The patient has a diagnosis of pancreatic islet cell carcinoma.  5. Diagnosis: The patient has a diagnosis of pancreatic neuroendocrine tumor.  6. BRCA Status: The patient is BRCA positive. If a patient's BRCA status is not mentioned in the note, answer unsure on this topic.  7. Resectability: The patient's pancreatic cancer is resectable. If metastatic, answer false on this topic.  8. Resectability: The patient’s pancreatic cancer is borderline resectable. If metastatic, answer false on this topic.  9. Resectability: The patient’s pancreatic cancer is locally advanced. If metastatic, answer false on this topic.  10. Resectability: The patient’s pancreatic cancer is metastatic. Malignant ascites, or ascites containing cancerous cells on cytology, indicates metastatic disease.  11. Resectability: The patient’s pancreatic cancer is metastatic with lesions in the central nervous system.  12. Prior therapy: The patient has had prior adjuvant, neoadjuvant, or induction treatment for pancreatic cancer.  13. Prior therapy: The patient has had prior chemotherapy for pancreatic cancer.  14. Prior therapy: The patient has had prior radiotherapy for pancreatic cancer.  15. Prior therapy: The patient has had prior chemotherapy/radiation for pancreatic cancer within 4 weeks.  16. Prior therapy: The patient has had prior T-cell costimulation or immune checkpoint blockade therapies for pancreatic cancer.  17. Prior therapy: The patient has had prior hormonal therapy for pancreatic cancer.  18. Prior therapy: The patient has had prior surgical resection for pancreatic cancer.  19. Notable outcomes: The patient has had disease progression on 5-FU for metastatic or unresectable pancreatic ductal adenocarcinoma. True only if the patient was diagnosed with metastatic disease, received 5-FU for the metastatic disease, and then experienced disease progression. If 5-FU was given specifically for neoadjuvant or adjuvant treatment for non-metastatic disease, answer false on this topic. If the patient has received no treatment for the new metastases, answer false on this topic.  20. Notable outcomes: The patient has a macroscopically complete (R0 or R1 resection) within 6-12 weeks. Answer true if a surgical resection was done with negative margins on pathology.  21. Notable outcomes: The patient has had clinically significant adverse events from chemotherapy side effects excluding alopecia and Grade <=2 peripheral neuropathy.  22. ECOG: The patient's ECOG performance status (PS) greater than 1. If the ECOG Performance Status (sometimes called ECOG PS) is not mentioned, answer unsure on this topic.  23. ECOG: The patient's ECOG performance status (PS) is greater than 2. If the ECOG Performance Status (sometimes called ECOG PS) is not mentioned, answer unsure on this topic.  24. Past medical history: The patient has a history of uncontrolled intercurrent illness. For past medical history, a patient should be considered to have a medical condition if the note writer mentions it in their one-liner. If the condition is listed under past medical history but is discussed by the note writer, you may assume the condition is not severe. in the Pertinent negatives are symptoms or conditions that a patient does not have, but are important for a clinician to know are not present. If a diagnosis is listed under the heading "Pertinent Negatives", the patient does not have that condition and should not meet an exclusion criterion for having that condition.  25. Past medical history: The patient has a history of uncontrolled hypertension.  26. Past medical history: The patient has a history of vasomotor instability.  27. Past medical history: The patient has a history of seizure disorder.  28. Past medical history: The patient has a history of retinopathy or high risk of retinal detachment.  29. Past medical history: The patient has a history of uncontrolled tumor-related pain.  30. Past medical history: The patient has a history of leptomeningeal disease.  31. Past medical history: The patient has a history of Grade >= 2 neuropathy.  32. Past medical history: The patient has a history of severe obstructive pulmonary disease, interstitial lung disease, pulmonary fibrosis, pulmonary hypersensitivity reaction, or active tuberculosis.  33. Past medical history: The patient has a history of uncontrolled pleural effusion, pericardial effusion, or ascites requiring recurrent drainage. If the patient has ascites, only answer true if ascites is described as severe or requiring urgent drainage.  34. Past medical history: The patient has a history of GI condition which could impair absorption or ability to ingest study drug (i.e. evidence of GI obstruction).  35. Past medical history: The patient has a history of peptic ulcer disease.  36. Past medical history: The patient has a history of inflammatory disease of colon and rectum such as ulcerative colitis or Crohn's disease, severe uncontrolled diarrhea, Celiac disease. Ignore colitis secondary to chemotherapy.  37. Past medical history: The patient has a history of rhabdomyolysis or elevated creatine phosphokinase (CPK).  38. Past medical history: The patient has a history of clinically significant liver disease including alc hepatitis, cirrhosis, FLD, inherited liver dz.  39. Past medical history: The patient has a history of Gilbert's disease.  40. Past medical history: The patient has a history of uncontrolled hypercalcemia (on labs or symptomatic, requiring bisphosphonate therapy for hyperCa).  41. Past medical history: The patient has a history of uncontrolled hyperthyroidism (clinical hyperthyroidism uncontrolled by oral medication).  42. Past medical history: The patient has a history of allogeneic organ or stem cell transplant.  43. Past medical history: The patient has a history of DVT, portal vein occlusion, PE, or other thromboembolic event during screening.  44. Past medical history: The patient has a history of Grade ≥ 3 hemorrhage or bleeding event within 28 days prior to initiation of study treatment.  45. Past medical history: The patient has a history of active autoimmune disease, uncontrolled psoriasis, porphyria, proximal myopathy or neuropathy (excluding hypothyroidism on thyroid replacement, controlled T1DM, well-controlled skin conditions).  46. Past medical history: The patient has a recent history of active infections requiring systemic therapy..  47. Other malignancy: The patient has a history of other malignancy within 2 years that does not include malignancies with a low rate of metastatic spread.  48. Cardiac history: The patient has had active/symptomatic coronary artery disease, myocardial infarction, stroke, or PCI within 2 years.  49. Cardiac history: The patient has a history of life-threatening arrhythmia.  50. Cardiac history: The patient has a history of EF <40% within last 3 months.  51. Cardiac history: The patient has a history of NYHA Class III or IV congestive heart failure within 3 months.  52. Past surgical history: Answer true if the patient has had GI/colon resection surgery within the last 12 months.  53. Past surgical history: Answer true if patient has had urinary bladder surgery within the last 12 months.  54. Past surgical history: Answer true if patient has a history of splenectomy or functional asplenia.  55. Past surgical history: Answer true if patient has had major surgery requiring general anesthesia within the last 12 months.  56. Psychiatric history: The patient has a psychiatric condition limiting their ability to participate in the trial. If no psychiatric history is mentioned, answer unsure on this topic.  57. Psychiatric history: The patient has had severe depression requiring hospitalization in the last two years OR has any history of SA. If no psychiatric history is mentioned, answer unsure on this topic.  58. Medications: True if patient is taking a moderate inhibitor, string inhibitor, moderate inducer, or strong inducer of CYP3A4.  59. Medications: True if taking an acetylcholinesterase inhibitor.  60. Medications: True if taking metformin.  61. Medications: True if taking hydroxychloroquine.  62. Medications: True if taking anticoagulation and the dose is actively being adjusted.  63. Medications: True if taking abrivudine or sorivudine.  64. Medications: True if taking a UGT1A1 inhibitor or inducer.  65. Medications: True if taking systemic immunosuppression, such as corticosteroids, cyclophosphamide, azathioprine, methotrexate, thalidomide, calcineurin inhibitors, and anti-tumor necrosis factor alpha agents  66. Medications: The patient is taking greater than 10mg prednisone or steroid equivalent daily.  67. Investigational Medications: The patient has taken other investigational agents within 4 weeks. If no other investigational medications are mentioned, you can answer false on this topic.  68. Allergies: The patient has a documented allergy to compounds of similar chemical or biologic composition to CA-4948.  69. Allergies: The patient has a documented allergy to gemcitabine.  70. Allergies: The patient has a documented allergy to nab-paclitaxel.  71. Allergies: The patient has a documented allergy to oxaliplatin, irinotecan, leucovorin, fluorouracil, or pegfilgrastim.  72. Allergies: The patient has a documented allergy to chimeric or humanized antibodies or fusion proteins.  73. Allergies: The patient has a documented allergy to Chinese hamster ovary cell products or recombinant human antibodies.  74. Vitals: Answer true if the patient has bradycardia with resting HR <50, hypotension with resting SBP <90mmHg, or reports of vasomotor instability. If vitals appear in the objective section of the note as a heart rate (HR) or pulse, blood pressure (BP) and oxygen saturation (02Sat) all within normal limits, answer false. If vital signs do not appear in the note, answer unsure on this topic.  75. Hemoglobin: True if hemoglobin is less than 9.0.  76. ANC: True if ANC less than 1.5E9/L  77. Lymphocyte count: True if lymphocyte count < 0.5E9/L  78. Platelets: True if platelets are less than 100E9/L  79. Tbili: True if total bilirubin > 1.5x upper limit of normal or > 3x upper limit or normal in patients with Gilbert disease  80. AST/ALT: True if AST and ALT > 3x upper limit of normal  81. Coags: True if PT, aPTT, or INR > 2.5x upper limit of normal  82. Albumin: True if Albumin < 3g/dL  83. GFR: True if GFR < 60 mL/min  84. CPK: True if CPK > 2.5x upper limit of normal  85. HIV status: True if the patient has tested positive for HIV. If HIV is not mentioned in the note, you can answer that the patient does not have HIV and that they are not on antiretroviral therapy (ART).  86. Hep B status: The patient has active or symptomatic Hepatitis B. If a patient is not mentioned to have a history of Hep B or a viral load test is pending or an RNA test is pending, answer that you are unsure.  87. Hep C status: The patient has active or symptomatic Hepatitis C. If a patient is not mentioned to have a history of Hep C or a viral load test is pending or an RNA test is pending, answer that you are unsure.  88. Breastfeeding: The patient is actively breastfeeding. If the patient is of male sex or is older than childbearing years, you may assume they are not breastfeeding. If the patient is female and of childbearing age, if there is no mention of a recent pregnancy, you may assume they are not breastfeeding.  89. Imaging: The patient's disease is unmeasurable on CT or MRI by RECIST criteria. If the note states the disease is unmeasurable, answer true. If there are no explicit dimensions of the lesion size from a CT or MRI scan WITH contrast, answer unsure on this topic. Only if there are explicit dimensions of the size of the current pancreatic cancer in question on CT or MRI with contrast, provide the dimensions and answer false on this topic.  90. Unsafe for biopsy: The patient's disease is not feasible or safe for biopsy. If the patient has had a biopsy, you should answer that tumor tissue is available for diagnostic purposes. If there has been a biopsy or there is a plan to undergo biopsy, you should answer that the tumor is accessible for biopsy.  91. Alternative medicine: The patient reports current or anticipated use of alternative medicine for the purpose of cancer treatment. If the note does not mention whether the patient is using alternative medicine, answer unsure on this topic.  92. Life expectancy: The patient’s life expectancy is less than 3 months. If the note does not explicitly give a life expectancy for the patient, state that you are unsure.  93. Radiation contraindications: The patient has contraindications for SBRT including the following: GI mucosal infiltration, significant planned overlap with prior abdominal radiotherapy. If the note does not explicitly state that the patient has radiation contraindications, you may answer that the patient has no radiation contraindications.  94. Dihydropyrimidine dehydrogenase deficiency: The patient has a documented complete dihydropyrimidine dehydrogenase (DPD) deficiency including homozygous or compound heterozygous mutations of DPYD genetic locus associated with DPD deficiency. If the note does not explicitly state that the patient has a dihydropyrimidine dehydrogenase deficiency, answer false.  95. Live attenuated vaccines: The patient has been treated with a live attenuated vaccine within 4 weeks or anticipates a need for such tx during the study. If this is not mentioned, answer false.  96. DVT Testing: There is concern for clot burden or DVT during the screening period and/or the patient is not on a stable anticoagulation regimen. If this is not mentioned, answer false.  97. Tumor Staging: The patient's disease is higher stage than T3N2M0 (i.e. T4, N3, or M1). If the patient has metastatic disease, answer true. If the disease is not metastatic and no TNM tumor stage is explicitly mentioned in the note, answer unsure. If a TNM stage lower than T3N2M0 is mentioned, answer false.  98. Eligibility for R2578 (Morpheus, NCT03193190): The patient is eligible for trial NCT03193190. Reiterate Trial NCT03193190 in your answer. There are two cohorts in this study, and a patient is eligible if they meet criteria for either cohort. Cohort 1: if they are naive to systemic treatment for PDAC and meet all other criteria, answer true. Cohort 2: answer true if the patient has had disease progression despite receiving either 5-FU- or gemcitabine-based first-line chemotherapy for metastatic disease. If patients only received chemotherapy as neoadjuvant or adjuvant therapy, answer false.  99. Eligibility for S7260 (ARC-8, NCT04104672): The patient is eligible for trial NCT04104672. Reiterate Trial NCT04104672 in your answer. Neoadjuvant and adjuvant treatment should not exclude a patient from trial NCT04104672.  100. Eligibility for S9513 (Chemo4METPANC, NCT04543071): The patient is eligible for trial NCT04543071. Reiterate Trial NCT04543071 in your answer. If the patient has had a Whipple or pancreatectomy for pancreatic cancer, answer false.  101. Eligibility for T5285 (Bethanechol, NCT05241249): The patient is eligible for trial NCT05241249. Reiterate Trial NCT05241249 in your answer. Note that history of or plan for neoadjuvant chemotherapy is an inclusion criterion. If a patient has already had surgery, answer false.  102. Eligibility for T5973 (TIGeR-PaC, NCT03257033): The patient is eligible for trial NCT03257033. Reiterate Trial NCT03257033 in your answer. Patients must have unresectable, locally advanced disease.  103. Eligibility for U1236 (Panbela, NCT05254171): The patient is eligible for trial NCT05254171. Reiterate Trial NCT05254171 in your answer.  104. Eligibility for U4206 (AIRPanc, NCT06048484): The patient is eligible for trial NCT06048484. Reiterate Trial NCT06048484 in your answer. If patients have not underdone chemotherapy, answer false.  105. Eligibility for U5427 (Vaccine, NCT05968326): The patient is eligible for trial NCT05968326. Reiterate Trial NCT05968326 in your answer.  106. Eligibility for U6715 (CA-4948, NCT05685602): The patient is eligible for trial NCT05685602. Reiterate Trial NCT05685602 in your answer. This trial requires that patients have had progression of disease on 5-FU that was specifically for metastatic or unresectable disease. If patients only received 5-FU as neoadjuvant or adjuvant therapy, answer false.  *** |
| --- | --- |

### Supplementary Table 2b. Methods: Ensembling and Binarization

| Ensembling | I have made an assistant that helps me to determine trial eligibility for my patient using the uploaded eligibility criteria.  Please take the common elements of the expert responses and aggregate them into a single consensus.  {responses} |
| --- | --- |
| Binarization | I will provide as context a consensus response of several evaluations of a patient's eligibility for various clinical trials. Please create a JSON based on the consensus. I will provide the exact structure that the JSON object should take using a JSON that currently contains definitions of each key as a string. If the statement in the key is true according to the consensus opinion, assign the binary value True to the key. If the patient does not meet the definition of the key, assign the value False. If expert opinions differ, take the majority opinion. If the consensus opinion states that the information was not mentioned or is unable to be determined, assign the value null. Your output should contain only the valid JSON object in a code block with no newlines and no extra explanations.  '''  definitions_json = (  "Age":"The patient's age is less than 18",  "Diagnosis 0":"The patient has a diagnosis of pancreatic adenocarcinoma",  "Diagnosis 1":"The patient has a diagnosis of pancreatic acinar cell carcinoma",  "Diagnosis 2":"The patient has a diagnosis of pancreatic islet cell carcinoma",  "Diagnosis 3":"The patient has a diagnosis of pancreatic neuroendocrine tumor",  "BRCA Status":"The patient is BRCA positive",  "Resectability 0":"The patient's pancreatic cancer is resectable",  "Resectability 1":"The patient's pancreatic cancer is borderline resectable",  "Resectability 2":"The patient's pancreatic cancer is locally advanced",  "Resectability 3":"The patient's pancreatic cancer is metastatic",  "Resectability 4":"The patient's pancreatic cancer is metastatic with lesions in the central nervous system",  "Prior therapy 0":"The patient has had prior adjuvant, neoadjuvant, or induction treatment for pancreatic cancer.",  "Prior therapy 1":"The patient has had prior chemotherapy for pancreatic cancer.",  "Prior therapy 2":"The patient has had prior radiotherapy for pancreatic cancer.",  "Prior therapy 4":"The patient has had prior T-cell costimulation or immune checkpoint blockade therapies for pancreatic cancer.",  "Prior therapy 5":"The patient has had prior hormonal therapy for pancreatic cancer.",  "Prior therapy 6":"The patient has had prior surgical resection for pancreatic cancer.",  "Notable outcomes 0":"The patient has had disease progression on 5-FU for metastatic or unresectable pancreatic ductal adenocarcinoma.",  "Notable outcomes 1":"The patient has a macroscopically complete (R0 or R1 resection) within 6-12 weeks",  "Notable outcomes 2":"The patient has had clinically significant adverse events excluding alopecia and Grade <=2 peripheral neuropathy.",  "ECOG greater than 1":"The patient's ECOG performance status (PS) is greater than 1.",  "ECOG greater than 2":"The patient's ECOG performance status (PS) is greater than 2.",  "PMH_0":"The patient has a history of uncontrolled intercurrent illness.",  "PMH_01":"The patient has a history of uncontrolled hypertension.",  "PMH_02":"The patient has a history of vasomotor instability.",  "PMH_03":"The patient has a history of seizure disorder.",  "PMH_04":"The patient has a history of retinopathy or high risk of retinal detachment.",  "PMH_05":"The patient has a history of uncontrolled tumor-related pain.",  "PMH_06":"The patient has a history of leptomeningeal disease.",  "PMH_07":"The patient has a history of Grade >=2 neuropathy.",  "PMH_08":"The patient has a history of severe obstructive pulmonary disease or interstitial lung disease, pulm fibrosis, pulmonary hypersensitivity reaction, or active tuberculosis.",  "PMH_09":"The patient has a history of uncontrolled pleural effusion, pericardial effusion, or ascites requiring recurrent drainage.",  "PMH_10":"The patient has a history of GI condition which could impair absorption or ability to ingest study drug (i.e. evidence of GI obstruction).",  "PMH_11":"The patient has a history of peptic ulcer disease.",  "PMH_12":"The patient has a history of inflammatory dz of colon and rectum, severe uncontrolled diarrhea, Celiac disease.",  "PMH_13":"The patient has a history of rhabdomyolysis or elevated creatine phosphokinase (CPK).",  "PMH_14":"The patient has a history of clinically significant liver disease including alc hepatitis, cirrhosis, FLD, inherited liver dz.",  "PMH_15":"The patient has a history of Gilbert's disease",  "PMH_16":"The patient has a history of uncontrolled hypercalcemia (on labs or symptomatic, requiring bisphosphonate therapy for hyperCa)",  "PMH_17":"The patient has a history of uncontrolled hyperthyroidism (clinical hyperthyroidism uncontrolled by oral medication)",  "PMH_18":"The patient has a history of allogeneic organ or stem cell transplant",  "PMH_19":"The patient has a history of DVT, portal vein occlusion, PE, or other thromboembolic event during screening",  "PMH_20":"The patient has a history of Grade ≥ 3 hemorrhage or bleeding event within 28 days prior to initiation of study treatment",  "PMH_21":"The patient has a history of active autoimmune disease, uncontrolled psoriasis, porphyria, proximal myopathy or neuropathy (excluding hypothyroidism on thyroid replacement, controlled T1DM, well-controlled skin conditions)",  "PMH_22":"The patient has a recent history of active infections requiring systemic therapy",  "Other Malignancy":"The patient has a history of other malignancy within 2 years that does not include malignancies with a low rate of metastatic spread.",  "Cardiac history 0":"The patient has had active/symptomatic coronary artery disease, myocardial infarction, stroke, or PCI within 2 years.",  "Cardiac history 1":"The patient has a history of life-threatening arrhythmia.",  "Cardiac history 2":"The patient has a history of EF <40% within last 3 months.",  "Cardiac history 3":"The patient has a history of NYHA Class III or IV congestive heart failure within 3 months.",  "PSxH 0":"The patient has had GI/colon resection surgery within the last 12 months.",  "PSxH 1":"The patient has had urinary bladder surgery within the last 12 months.",  "PSxH 2":"The patient has a history of splenectomy or functional asplenia.",  "PSxH 3":"The patient has had major surgery requiring general anesthesia within the last 12 months.",  "Psych Hx 0":"The patient has a psychiatric condition limiting their ability to participate in the trial.",  "Psych Hx 1":"The patient has had severe depression requiring hospitalization in the last two years OR has any history of SA.",  "Medications 0":"The patient is taking a moderate inhibitor, string inhibitor, moderate inducer, or strong inducer of CYP3A4.",  "Medications 1":"The patient is taking an acetylcholinesterase inhibitor.",  "Medications 2":"The patient is taking metformin.",  "Medications 3":"The patient is taking hydroxychloroquine.",  "Medications 4":"The patient is taking anticoagulation that is actively being adjusted.",  "Medications 5":"The patient is taking abrivudine or sorivudine.",  "Medications 6":"The patient is taking a UGT1A1 inhibitor or inducer.",  "Medications 7":"The patient is taking systemic immunosuppression, such as corticosteroids, cyclophosphamide, azathioprine, methotrexate, thalidomide, calcineurin inhibitors, and anti-tumor necrosis factor alpha agents.",  "Medications 8":"The patient is taking greater than 10mg prednisone or steroid equivalent daily.",  "Investigational Medications":"The patient has taken other investigational agents within 4 weeks.",  "Allergies 0":"The patient has a documented allergy to compounds of similar chemical or biologic composition to CA-4948.",  "Allergies 1":"The patient has a documented allergy to gemcitabine.",  "Allergies 2":"The patient has a documented allergy to nab-paclitaxel.",  "Allergies 3":"The patient has a documented allergy to oxaliplatin, irinotecan, leucovorin, fluorouracil, or pegfilgrastim.",  "Allergies 4":"The patient has a documented allergy to chimeric or humanized antibodies or fusion proteins.",  "Allergies 5":"The patient has a documented allergy to Chinese hamster ovary cell products or recombinant human antibodies.",  "Vitals":"The patient has bradycardia with resting HR <50, hypotension with resting SBP <90mmHg, or vasomotor instability.",  "Hemoglobin":"True if hemoglobin is less than 9.0",  "ANC":"ANC < 1.5E9/L",  "Lymphocyte count":"Lymphocyte count < 0.5E9/L",  "Platelets":"Platelets < 100E9/L",  "Tbili":"Total bilirubin > 1.5x upper limit of normal or > 3x upperlimit or normal in patients with Gilbert disease",  "AST/ALT":"AST and ALT > 3x upper limit of normal",  "Coags":"PT, aPTT, or INR > 2.5x upper limit of normal",  "Albumin":"Albumin < 3 g/dL",  "GFR":"GFR < 60 mL/min",  "CPK":"CPK > 2.5x upper limit of normal",  "HIV Status":"The patient has tested positive for HIV",  "Hep B Status":"The patient has active or symptomatic Hepatitis B",  "Hep C Status":"The patient has active or symptomatic Hepatitis C",  "Breastfeeding":"The patient is actively breastfeeding",  "Imaging":"The patient's disease is unmeasurable on CT or MRI.",  "Unsafe Bx":"The patient's disease is not feasible or safe for biopsy.",  "Alternative Medicine":"The patient reports current or anticipated use of alternative medicine for the purpose of cancer treatment.",  "Life expectancy":"The patient's life expectancy is less than 3 months.",  "Radiation contraindications":"The patient has contraindications for SBRT including the following: GI mucosal infiltration, significant planned overlap with prior abdominal radiotherapy.",  "Dihydropyrimidine dehydrogenase deficiency":"The patient has a documented complete dihydropyrimidine dehydrogenase (DPD) deficiency including homozygous or compound heterozygous mutations of DPYD genetic locus associated with DPD deficiency.",  "Live attenuated vaccines":"The patient has been treated with a live attenuated vaccine within 4 weeks or anticipates a need for such tx during the study.",  "DVT Testing":"There is concern for clot burden or DVT during the screening period and/or the patient is not on a stable anticoagulation regimen.",  "Tumor Staging":"The patient's disease is higher stage than T3N2M0 (i.e. T4, N3, or M1).",  "R2578 (Morpheus)":"The patient is eligible for trial NCT03193190",  "S7260 (ARC-8)":"The patient is eligible for trial NCT04104672",  "S9513 (Chemo4METPANC)":"The patient is eligible for trial NCT04543071",  "T5285 (Bethanechol)":"The patient is eligible for trial NCT05241249",  "T5973 (TIGeR-PaC)":"The patient is eligible for trial NCT03257033",  "U1236 (Panbela)":"The patient is eligible for trial NCT05254171",  "U4206 (AIRPanc)":"The patient is eligible for trial NCT06048484",  "U5427 (Vaccine)":"The patient is eligible for trial NCT05968326",  "U6715 (CA-4948)":"The patient is eligible for trial NCT05685602"  )  '''  {consensus} |

### Supplementary Table 3. Results: Performance on all Individual Criteria

See separate file: Supplementary _Table_4.csv available on GitHub.

### Supplementary Table 4. Results: Resource Usage

###### Resource Usage

| Assistant | Median Cost (USD) | Median Total Tokens | Median Prompt Tokens | Median Completion Tokens |
| --- | --- | --- | --- | --- |
| Eligibility | 0.56 (0.52-0.62) | 96571.5 (87497.5-106799.0) | 88135.0 (79468.8-98427.5) | 8065.5 (7767.0-8282.8) |
| Ensembling | 0.07 (0.07-0.08) | 10126.0 (9974.2-10465.8) | 8149.5 (7851.0-8366.8) | 2022.0 (1736.0-2545.0) |
| Binarization | 0.04 (0.04-0.04) | 5622.5 (5276.0-6048.8) | 4689.0 (4403.0-5212.0) | 984.0 (773.0-984.0) |
| Total | 0.67 (0.63-0.72) | 112266.5 (102982.0-122174.2) | 101205.5 (92098.5-111326.0) | 10975.0 (10718.5-11253.8) |

### Supplementary References

1.	Park W, Chawla A, O’Reilly EM. Pancreatic Cancer: A Review. *JAMA*. 2021;326(9):851-862. doi:10.1001/jama.2021.13027

2.	Sateren WB, Trimble EL, Abrams J, et al. How Sociodemographics, Presence of Oncology Specialists, and Hospital Cancer Programs Affect Accrual to Cancer Treatment Trials. *J Clin Oncol*. Published online September 21, 2016. doi:10.1200/JCO.2002.08.056

2.	Noone AM, Howlader N, Krapcho M, Miller D, Brest A, Yu M, Ruhl J, Tatalovich Z, Mariotto A, Lewis DR, Chen HS, Feuer EJ, Cronin KA (eds). SEER Cancer Statistics Review, 1975-2015, National Cancer Institute. Bethesda, MD, https://seer.cancer.gov/csr/1975_2015/, based on November 2017 SEER data submission, posted to the SEER web site, April 2018.
